# Chronic Coronary Syndrome in Mexico: Design and Initial Insights from the RESINCCRO Mexican Registry

**DOI:** 10.64898/2026.07.15.26358091

**Authors:** Enrique Alexander Berrios-Bárcenas, Manuel Odín de los Ríos-Ibarra, Marco Antonio Alcocer-Gamba, Carlos Ramon Rodas-Cáceres, Edith Dalilia Ruiz-Gastelum, Manuel Alfonso Baños-González, Estrella Milbeth Vizarraga-Thomas, Martin de J. Valenzuela-Valenzuela, Francisco Gerardo Padilla-Padilla, Luis Gerardo González-Barrera, Juan Manuel Rebull-Isusi, Arcenio Alfonso Lendo-López, Alberto Esteban Bazzoni-Ruiz, Francisco Javier Roldán-Gómez, Héctor González-Godínez, Carlos Hernández-Herrera, M. Cecilia Escalante-Seyffert, Juan Pablo Núñez-Urquiza, José Luis Leiva-Pons, Juan Rene Cornejo-Avendaño, Erika Danahe Duarte-Montiel, Alejandra Portillo-Romero, Patricia L. Nuriulu-Escobar, Raúl Navarrete-Gaona, Humberto Rodríguez-Reyes, Manuel R. Barrera-Bustillos, José Salvador Laínez-Zelaya, Alejandro Ricalde-Alcocer, Alejo Diaz-Aragón, Jorge Cortés-Lawrenz, Sergio A. Chávez-Leal, Francisco Cruz-Ramos, Yuri Bastien-Araujo, Juan José Cruz-Cruz, Hipólito Alfredo Pérez-Sandoval, Norberto Matadamas-Hernández, Mauricio Rivera-Vela, Jorge Abel Vázquez-Acosta, Abelardo Burgueño-Rivas, Luisa F. Aguilera-Mora, Sergio Arturo Flores-Velasco, Ángel Iván González-Pacheco, Gerardo Santiago Baca-Escobar, Juan José Parcero-Valdés, José Arturo Maldonado-Villalon, Erick Alexanderson-Rosas, Alexandra Arias-Mendoza, Abel Alberto Pavía-López, Francisco M. Baranda-Tovar, Carlos A. Guillen-Ortiz, the RESINCCRO group of study

## Abstract

**BACKGROUND:** Chronic coronary syndromes (CCS) remain under-characterized in Latin America, where clinical profiles may differ from high-income countries.

**OBJECTIVE:** We aim to characterize the clinical presentation, coronary anatomic profile, and pharmacologic treatment patterns of adults living with CCS using data from the Mexican Chronic Coronary Syndrome Registry (RESINCCRO).

**METHODS:** RESINCCRO is an observational, multicenter, cross-sectional registry conducted across ∼50 centers in five regions from Mexico. We included adults (≥18 years) enrolled between September 2024 and March 2025 who met 2019 ESC CCS criteria. Coronary imaging data was collected from medical records into a standardized electronic case report form.

**RESULTS:** We enrolled 3,029 adults (men [72.5%]; mean age 67.2□±□10.7 years). Cardiometabolic comorbidities were frequent: overweight/obesity (76%), arterial hypertension (69.0%), type 2 diabetes (44.0%), and chronic kidney disease (24.2%). Persistent angina/equivalents occurred in (23.9%), of which most had Canadian Cardiovascular Society class I–II (91.2%). The mean LVEF was of 53.7%□±□12.0. Cardiac rehabilitation participation was (6.2%). Median LDL-C was 70 mg/dL (IQR 51–95) and LDL <55 mg/dL was only 26.1%, despite high prescription of lipid-lowering therapies, including statins (93.2%), ezetimibe (24.6%), and PCSK9 inhibitors (2.4%). 60.3% had obstructive epicardial disease.

**CONCLUSIONS:** Mexican adults with CCS exhibit high cardiometabolic burden, frequent symptoms, suboptimal LDL-C goal attainment, low rehabilitation uptake, and a substantial obstructive phenotype. These findings highlight opportunities to intensify secondary prevention, adopt mechanism-directed evaluation and therapy, and expand cardiac rehabilitation to improve CCS care in Mexico.

Graphical abstract:
Chronic Coronary Syndrome in Mexico: design and initial insights from the RESINCCRO Mexican Registry. *Abbreviations*: CCS, chronic coronary syndromes; CCS class, Canadian Cardiovascular Society angina class; CKD, chronic kidney disease; CR, cardiac rehabilitation; ESC, European Society of Cardiology; IQR, interquartile range; LDL-C, low-density lipoprotein cholesterol; LVEF, left ventricular ejection fraction; OMT, optimal medical therapy; PCSK9, proprotein convertase subtilisin/kexin type 9; T2D, type 2 diabetes.

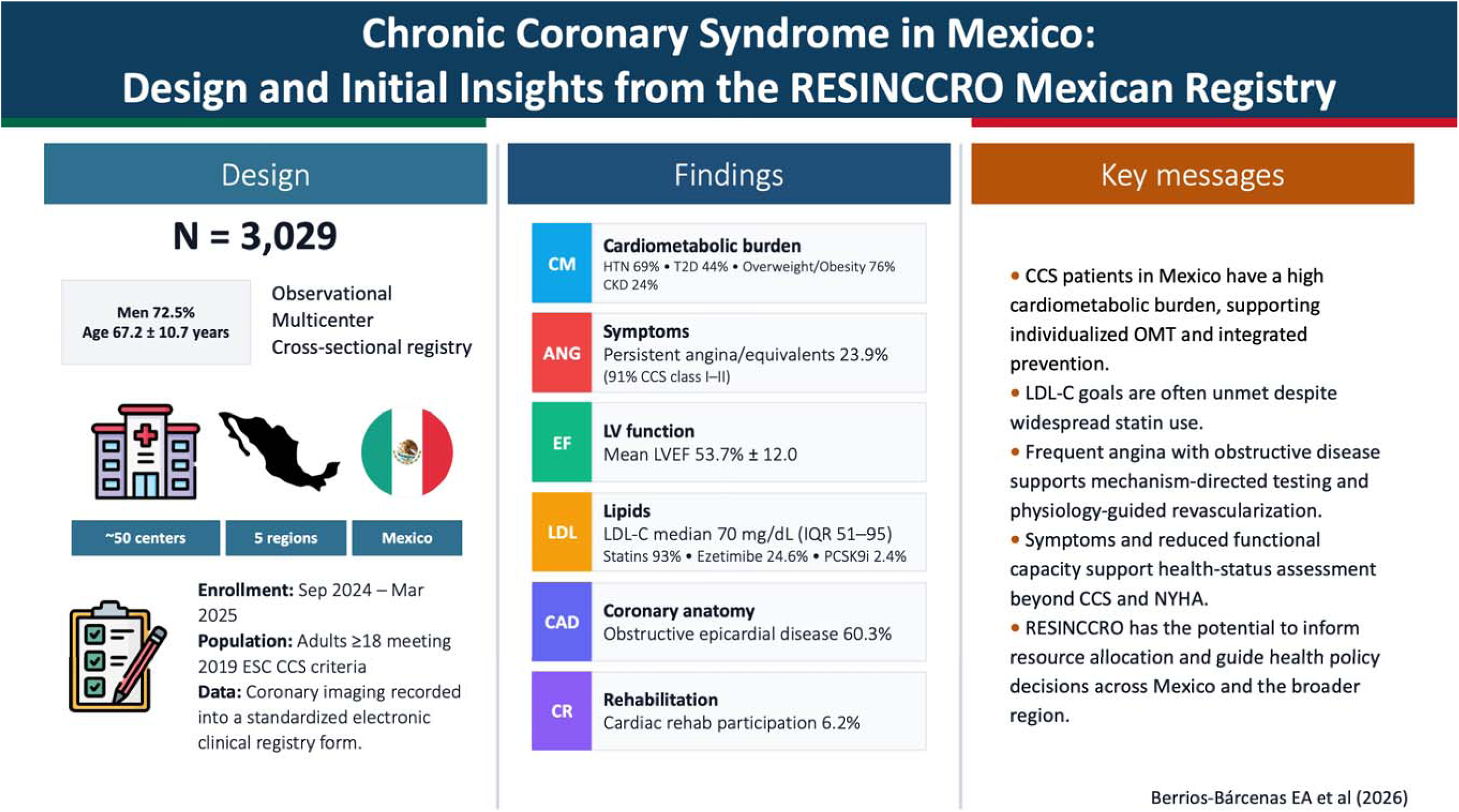

## INTRODUCTION

Coronary artery disease (CAD) is a pro-inflammatory process driven by endothelial dysfunction, atherosclerotic plaque deposition, and cellular activation within the microvascular circulation and epicardial coronary arteries. This pathophysiological process can lead to metabolic remodeling and coronary circulatory dysfunction, often accompanied by angina (1,2). CAD has been recognized as a cause of cardiovascular events, including adverse quality of life, as well as incident non-fatal myocardial infarction and cardiovascular mortality (3–5). Although prompt lifestyle change and pharmacologic or interventional therapies are often required, and can stabilize or even regress disease activity, recent evidence suggests that CAD can be best conceptualized as a chronic, progressive condition punctuated by episodic events rather than a “stable” state (6).

Within this spectrum, CAD is classified into acute coronary syndromes (ACS) and chronic coronary syndromes (CCS), both commonly presenting with exertional or rest angina as the symptomatic manifestation of myocardial supply–demand mismatch arising from obstructive, thrombotic, microvascular, or vasospastic mechanisms (1,7). Notably, hemodynamically significant epicardial stenosis may be absent in a substantial proportion of patients with angina or documented ischemia, underscoring the heterogeneity of CCS (6–8). To better capture this heterogeneity, in 2019 the European Society of Cardiology (ESC) issued clinical guidelines that classify CCS into six clinical categories spanning symptomatic suspicion of CAD, new heart failure with suspected ischemic etiology, post-event or post-revascularization states (<1 year and ≥1 year), vasospastic/microvascular angina, and asymptomatic individuals with CAD detected on screening (9). Other frameworks, such as the myocardial ischemic syndromes have been also proposed in order to better comprehend the clinical outcomes related to CCS (10).

However, from a population and epidemiologic perspective, CCS remains understudied and underrepresented across low- and middle-income countries, particularly in Latin America. This is particularly relevant for Latino populations, in whom CCS has often been described as having different risk-factor profiles, clinical features, and treatment patterns compared with high-income countries (11,12).

Although registries from Spain (e.g., PANES, REGICOR) and studies in U.S. Latino populations (e.g., NHLBI Dynamic Registry) have provide useful data, they may underrepresent local estimates in Latino populations with distinct risk profiles and health-care access (13–15). In Mexico, national registries such as the RENASICA I, II and III have helped address the burden and care pathways of ACS; however, contemporary and comprehensive evidence for CCS remains limited (16–18).

To address this area of opportunity, we conceptualized and implemented the Chronic Coronary Syndromes Mexican Registry (Acronym in Spanish—RESINCCRO: REgistro de SÍNdromes Coronarios CRÓnicos) to systematically characterize CCS in real-world practice across participating centers in Mexico. RESINCCRO was designed to generate local, granular and regional evidence on the current situation of CCS from recent years. Ultimately, RESINCCRO could help clinicians and policymakers inform future risk-stratification algorithms, specific clinical care pathways, and health-policy planning for a large segment of patients living with CCS in Mexico. Furthermore, RESINCCRO could inform the situation of CCS across Latin American populations and serve as an example for country-level registries throughout the region, ultimately enabling to Latin American CCS registries.

Hence, in this study, we aim to provide initial insights from the RESINCCRO registry through two primary objectives: (1) characterize the clinical presentation, cardiovascular risk factors, and anatomic presentation of CCS; and (2) describe the pharmacologic treatment profiles of the included patients in the registry.

## METHODS

### Study design

RESINCCRO is an observational, cross-sectional, multicenter registry of ∼50 research centers distributed across five Mexican regions (north, central-west, metropolitan area, south, and central). The list of participating centers by region and their corresponding state is provided in **Supplementary Table S1**. Investigators at each site collected and pooled data from patients and medical records into a pre-designed electronic case report form on a secure, virtual platform (https://consam.mx/resinccro/login.aspx). Records are stored locally and then retrieved centrally for data management and analysis. No study-mandated interventional tests were subsequently performed. All data were anonymized, and each patient received a unique study identifier. A research investigator (MORI) assessed the validity of each record, with subsequent random sampling of cases to evaluate the accuracy of the information provided by each site. For this first report, we describe the characteristics of the patients enrolled from September 2024 to March 2025. We included in RESINCCRO adults (≥18 years) of any sex who provided consent and met ≥1 of the following criteria for CCS: (i) stable angina (≤10-minute chest discomfort with ≥1 typical feature, exertional, typical location/radiation, relieved by rest or nitrates in <5 minutes); (ii) documented ischemia (angina equivalents) on non-invasive stress testing; (iii) prior CCS diagnosis based on clinical evaluation and/or compatible non-invasive or invasive imaging aligned with ESC criteria; (iv) anatomically obstructive CAD on imaging (>50% stenosis or a Fractional Flow Reserve [FFR] ≤0.80); (v) local expert diagnosis of ischemic heart disease/CAD; or (vi) signs/symptoms of heart failure plus ≥1 of the above (9). Although the ESC endorses in 2024 a novel classification for CCS, RESINCCRO was designed in 2021 where this approach was integrated for the creation of CCS registries. In **Supplementary Table S2**, we provide a full description and definitions of these eligibility criteria. We did not include individuals with acute coronary syndrome within the previous 2 months, unstable angina within 3 months, severe valvular disease, storage/infiltrative cardiomyopathy, hypertrophic/dilated/hereditary cardiomyopathies, aortic disease, pericardial disease or myocarditis, stress-induced cardiomyopathy, pulmonary hypertension, cardiac tumors, congenital heart disease, active or terminal cancer, or a positive pregnancy test. We also did not include patients who withdrew consent or had incomplete medical records. All patients provided written consent for their medical information to be included in RESINCCRO. The protocol was approved by the Research and Ethics Committee of Medica Sur, S.A.B. de C.V. (Protocol No. CEI-000005 – IC4-06795-011-MEX; **Supplementary Figure S1**). The study adheres to the principles of the Declaration of Helsinki and the reporting of the study to the Strengthening the Reporting of Observational Studies in Epidemiology (STROBE) guidelines for cross-sectional studies (**Supplementary Table S3**) (19).

### Assessed variables

RESINCCRO was designed to capture key CCS variables, which included the following domains: sociodemographic variables, cardiovascular risk factors and cardiovascular concomitant comorbidities, clinical presentation of CCS, vital signs and laboratory values, imaging studies and anatomic characterization of CAD and concomitant treatments (medical and revascularization). A concise overview of the included variables per each domain is presented in **Table 1**, and the extended variable catalogue is presented in **Supplementary Table S4**.

**Table 1:** Data collected in the RESINCCRO registry.

| Socioeconomic variables | Cardiovascular risk factors and cardiovascular comorbidities | Clinical presentation of CCS and vital signs and laboratory values | Imaging studies, procedures, and medication use |
| --- | --- | --- | --- |
| Age<br>Sex<br>Ethnic origin<br>Country of birth<br>Place of residence<br>Health-care coverage | **Cardiovascular risk factors:**<br>BMI and BMI categories<br>Arterial hypertension<br>Type 2 diabetes and microvascular complications<br>Thyroid dysfunction<br>CKD stage (G1–G5)<br>COPD<br>Anemia<br>Chronic liver disease<br>GERD<br>Osteoporosis<br>Smoking<br>Alcohol use disorder<br>**Cardiovascular comorbidities:**<br>Coronary artery disease<br>Peripheral arterial disease<br>Carotid artery disease<br>Stroke<br>Prior stroke/TIA<br>Atrial fibrillation<br>Sinus node dysfunction<br>Venous thromboembolism | **Clinical presentation of CCS**<br>Persistent angina/equivalent<br>CCS angina class (I–IV)<br>Maximum daily activity (I–IV)<br>**Vital signs and laboratory values**<br>Heart rate (bpm)<br>SBP (mmHg)<br>DBP (mmHg)<br>Hemoglobin (g/dL)<br>Platelets ($\times 10^9/L$)<br>Creatinine (mg/dL)<br>Glucose (mg/dL)<br>Urea (mg/dL)<br>Total cholesterol (mg/dL)<br>LDL-C (mg/dL)<br>HDL-C (mg/dL)<br>Triglycerides (mg/dL)<br>BNP (pg/mL)<br>NT-proBNP (pg/mL)<br>TSH ($\mu IU/mL$)<br>eGFR (CKD-EPI; mL/min/1.73 m<sup>2</sup>) | **Imaging studies and anatomic characterization of CAD**<br>Obstructive CAD<br>High-risk CAD<br>Vessels affected<br>Number of diseased vessels<br>CTO<br>LVEF<br>**Procedures**<br>Prior PCI<br>Prior CABG<br>Number of stents placed<br>**Medications (current/previous):**<br>Aspirin<br>Clopidogrel<br>Prasugrel<br>Ticagrelor<br>DOACs<br>Beta-blockers<br>ACE inhibitors<br>ARBs<br>Statins<br>Ezetimibe<br>PCSK9 inhibitors<br>Nitrates<br>Trimetazidine<br>Ivabradine |

*Abbreviations:* CE, angiotensin-converting enzyme; ARB, angiotensin II receptor blocker; BMI, body mass index; CABG, coronary artery bypass grafting; CAD, coronary artery disease; CCS, Chronic Coronary Syndrome; CCS class, Canadian Cardiovascular Society angina class; CKD, chronic kidney disease; COPD, chronic obstructive pulmonary disease; CTO, chronic total occlusion; DOAC, direct oral anticoagulant; GERD, gastroesophageal reflux disease; LVEF, left ventricular ejection fraction; PCI, percutaneous coronary intervention; PCSK9, proprotein convertase subtilisin/kexin type 9; TIA, transient ischemic attack.

### Statistical Analyses

A precision-based sample size of 2,967 (1,337 women; 1,630 men) was planned using the single-proportion Cochran formula, assuming CCS prevalences of 3.5% in women and 4.3% in men. (20) (21). Complete sample-size estimation is presented in the **Supplementary Methods**. Continuous variables were tested for distribution using the Kolmogorov–Smirnov test. Normally distributed variables are reported as mean (standard deviation, SD) and non-normal variables as median (interquartile range, IQR). Categorical variables are presented as frequencies and percentages. We report field-level completeness for all variables (n and % complete) in **Supplementary Table S4**. In the tables, we report missing values where applicable and provide the corresponding percentages when appropriate. No imputation was prespecified. For these first cross-sectional analyses, no formal multivariable modeling or inferential hypothesis testing was planned. All statistical analyses were performed using STATA (version 19). Figures were created using R Studio (Version 4.3.2) using the *ggplot2* R package (22).

## RESULTS

RESINCCRO collected data from 3,029 adults with CCS across the study period. Sociodemographic characteristics, cardiovascular risk factors and concomitant comorbidities are summarized in **Table 2**. Briefly, patients were predominantly male (72.5%) with a mean age of 67.2 ± 10.7 years. Most self-identified as Latino (93.6%), were born in Mexico (98.5%) and lived in urban areas (91.5%). The inclusion of the patients was mainly from Northern states (32.5%) and from Mexico City (24.2%) (**Figure 1**). Health-care coverage was split between the public (39.3%) and private (38.9%) sectors, with 21.8% reporting no coverage. Anthropometry showed a median BMI of 27.9 kg/m² (IQR: 25.2–30.8), with 44.8% classified as overweight and 23.7% in class I obesity. Regarding concomitant comorbidities, 69% of the sample had arterial hypertension, followed by type 2 diabetes (T2D) in 44%, and CKD in 24.2%. Among patients living with T2D, 7.1% had a history of diabetic nephropathy, 7.3% of diabetic retinopathy, and 13.1% of diabetic neuropathy. Regarding lifestyle habits, 33.7% were former smokers >1 year and 9.0% were classified as current smokers, while 2–7 drinks/week of alcohol intake was reported in 11.8%. Regarding cardiovascular history, 90% had a previous history of CAD, 4.0% of PAD, and 2.3% of carotid disease (**Figure 2**). Atrial fibrillation was recorded in 6.7%, 40.4% of whom had resting HR > 75 bpm.

**Figure 1:**
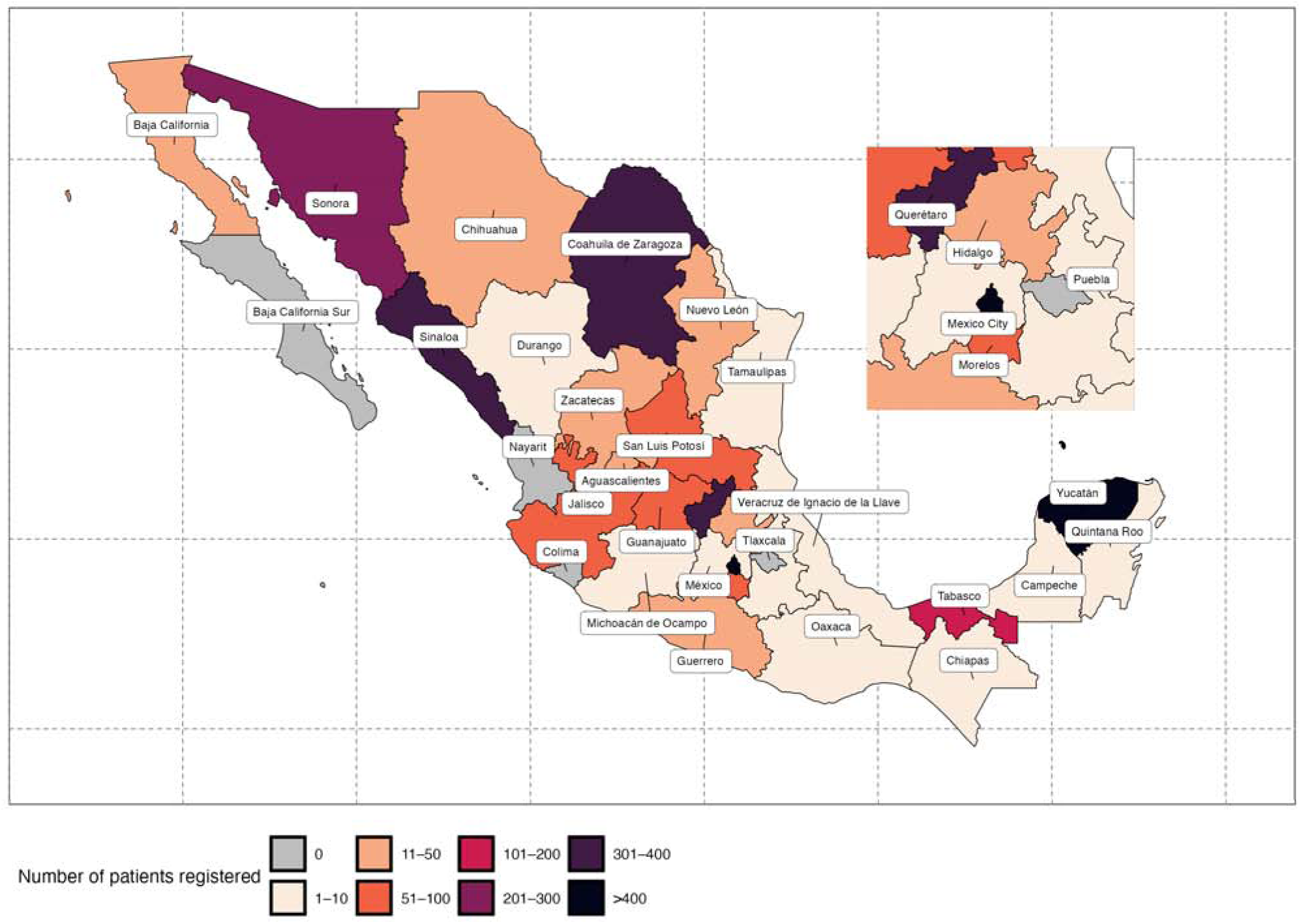
Geographic distribution of registered patients by state in the RESINCCRO registry. Data are shown as absolute counts (n) aggregated by state. The inset highlights the Metropolitan Area (Ciudad de México, Estado de México, Hidalgo, Morelos, Puebla, and Querétaro). *Abbreviations*: CDMX, Ciudad de México (Mexico City).

**Figure 2:**
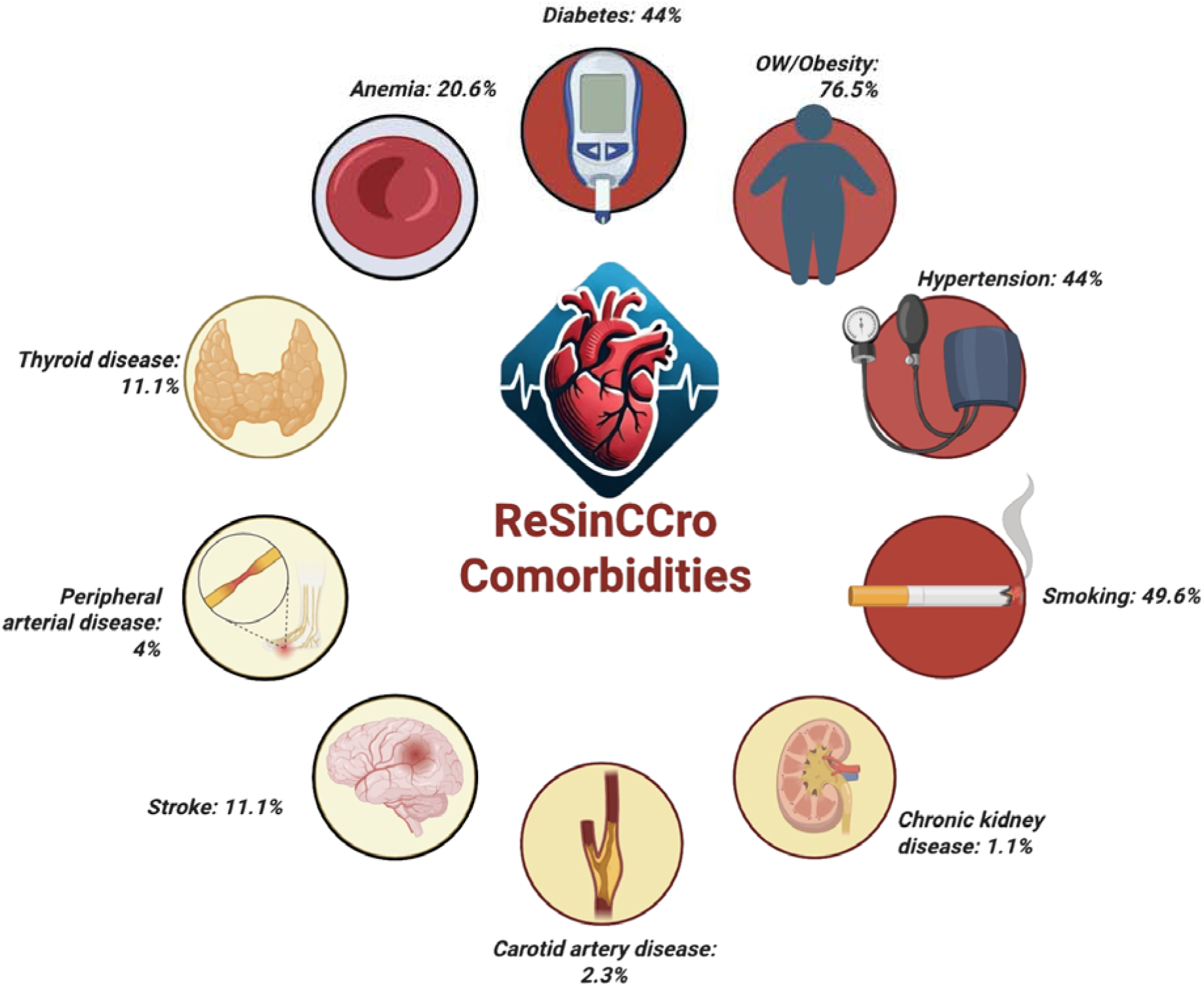
Cardiometabolic comorbidities in the 3,029 patients registered in the RESINCCRO. *Abbreviations*: RESINCCRO, REgistro de SÍNdromes Coronarios CRÓnicos; OW, overweight.

**Table 2:** Sociodemographic characteristics, cardiovascular risk factors and cardiovascular comorbidities of the patients included in RESINCCRO.

| Characteristics | n = 3,029 |
| --- | --- |
| *Demographic and socioeconomic profile* |  |
| **Age, (years)** | 67.2 ± 10.7 |
| **Sex, n (%)** |  |
| Male | 2,195 (72.5%) |
| Female | 834 (27.5%) |
| **Ethnic origin, n (%)** |  |
| Afro-descendant | 11 (0.4%) |
| White | 162 (5.3%) |
| Latino | 2,835 (93.6%) |
| Other | 21 (0.7%) |
| **Country of birth, n (%)** |  |
| Mexico | 2,984 (98.5%) |
| Foreign | 45 (1.5%) |
| **Place of residence, n (%)** |  |
| Urban | 2,772 (91.5%) |
| Rural | 257 (8.5%) |
| **Region of residence, n (%)** |  |
| North <sup>#</sup> | 985 (32.5%) |
| Central West <sup>#</sup> | 570 (18.8%) |
| Mexico City <sup>#</sup> | 732 (24.2%) |
| South <sup>#</sup> | 633 (20.9%) |
| Central <sup>#</sup> | 109 (3.6%) |
| **Health-care coverage, n (%)** |  |
| Public | 1,191 (39.3%) |
| Private | 1,177 (38.9%) |
| None | 661 (21.8%) |
| *Cardiovascular risk factors* |  |
| **BMI (kg/m<sup>2</sup>)** | 27.9 (25.2, 30.8) |
| **BMI categories, n (%)** |  |
| Underweight | 23 (0.8%) |
| Normal | 690 (22.8%) |
| Overweight | 1,357 (44.8%) |
| Obesity class I | 717 (23.7%) |
| Obesity class II | 187 (6.2%) |
| Obesity class III | 55 (1.8%) |
| **Arterial hypertension, n (%)** | 2,100 (69%) |
| **Type 2 diabetes, n (%)** | 1,334 (44%) |
| Nephropathy <sup>†</sup> | 95 (7.12%) |
| Retinopathy <sup>†</sup> | 97 (7.27%) |
| Neuropathy <sup>†</sup> | 175 (13.11%) |
| **Thyroid dysfunction, n (%)** |  |
| Hypothyroidism | 321 (10.6%) |
| Hyperthyroidism | 14 (0.5%) |
| **Chronic kidney disease, n (%)** | 733 (24.2%) |
| **eGFR categories, n (%)** |  |
| G1 | 759 (25.1%) |
| G2 | 1,418 (46.8%) |
| G3a | 398 (13.1%) |
| G3b | 195 (6.4%) |
| G4 | 89 (2.9%) |
| G5 | 51 (1.7%) |
| Missing | 121 (4%) |
| **COPD, n (%)** | 84 (2.8%) |
| **Anemia, n (%)** | 625 (20.6%) |
| **Chronic liver disease, n (%)** | 31 (1.0%) |
| **GERD, n (%)** | 193 (6.4%) |
| **Osteoporosis, n (%)** | 64 (2.1%) |
| **Smoking, n (%)** |  |
| Never | 1,527 (50.4%) |
| Former > 1 year | 1,022 (33.7%) |
| Former < 1 year | 107 (3.5%) |
| Passive | 29 (1.0%) |
| Active | 274 (9.0%) |
| Missing | 73 (2.4%) |
| **Alcohol intake, n (%)** |  |
| Never | 1,175 (38.8%) |
| > 1 year abstinent | 664 (21.9%) |
| < 1 drink/week | 560 (18.5%) |
| 2–7 drinks/week | 358 (11.8%) |
| 8–14 drinks/week | 66 (2.2%) |
| ≥ 15 drinks/week | 29 (1.0%) |
| Missing | 176 (5.8%) |
| *Cardiovascular comorbidities* |  |
| **History of CAD, n (%)** | 2,729 (90%) |
| **PAD, n (%)** | 124 (4.0%) |
| **Carotid artery disease, n (%)** | 70 (2.3%) |
| **Stroke, n (%)** | 112 (3.7%) |
| Ischemic <sup>§</sup> | 104 (92.9%) |
| Hemorrhagic <sup>§</sup> | 8 (7.1%) |
| **Atrial fibrillation, n (%)** | 203 (6.7%) |
| AF with resting HR > 75 bpm <sup>‡</sup> | 82 (40.4%) |
| **Sinus node dysfunction, n (%)** | 172 (5.7%) |
| **Venous thromboembolism, n (%)** | 33 (1.1%) |

Data are shown as mean ± SD, median [IQR], or n (%). **Annotations:** # = Regions were defined as follows: North: Baja California, Baja California Sur, Sonora, Sinaloa, Chihuahua, Coahuila, Nuevo León, Tamaulipas, Durango; Central West: Aguascalientes, Zacatecas, San Luis Potosí, Guanajuato, Querétaro, Jalisco, Colima, Michoacán, Nayarit; Central: Estado de México, Hidalgo, Morelos, Puebla, Tlaxcala; South: Guerrero, Oaxaca, Chiapas, Veracruz, Tabasco, Campeche, Yucatán, Quintana Roo. † = Percent among patients with type 2 diabetes; ‡ = Percent among patients with atrial fibrillation; § = Percent among patients with stroke. **Abbreviations:** AF, atrial fibrillation; BMI, body mass index; CAD, coronary artery disease; CKD, chronic kidney disease; COPD, chronic obstructive pulmonary disease; DBP, diastolic blood pressure; eGFR, estimated glomerular filtration rate; GERD, gastroesophageal reflux disease; HR, heart rate; IQR, interquartile range; PAD, peripheral arterial disease; SBP, systolic blood pressure

### Clinical presentation of CCS, vital signs, and laboratory assessment

In **Table 3**, we summarized clinical presentation, imaging findings, and procedures of the studied sample. Briefly, persistent angina or an angina equivalent was present in 23.9%, and 6.2% had undertaken cardiac rehabilitation. Angina burden assessed with the Canadian Cardiovascular Society criteria showed a trend toward lower classes, with class I the most prevalent (57.3%), followed by class II (33.9%), class III (7.2%), and class IV (1.6%). Functional capacity was predominantly sedentary (36.7%) or light (32.5%), while 29.4% reported moderate and 1.4% vigorous daily activity. Regarding vital signs, the average SBP was 124.8 ± 18.6 mmHg and the DBP 72.3 ± 10.1 mmHg. For the laboratory assessment, the median creatinine was 0.96 mg/dL [IQR: 0.80–1.17], glucose 104 mg/dL [IQR: 94–122], LDL-C 70 mg/dL [IQR: 51–95], HDL-C 42 mg/dL [IQR: 36–51], and triglycerides 123 mg/dL [IQR: 90–165], with a mean eGFR of 73.8 ± 23.0 mL/min/1.73 m². Only 26.1% (n= 792) patients had an LDL <55 mg/dL. Among those with natriuretic peptides measurement (n=178), they demonstrated a modestly elevated median BNP (86.5 pg/mL [IQR: 40.7–262.5]) and NT-proBNP (136 pg/mL [IQR: 60–435]).

**Table 3:**
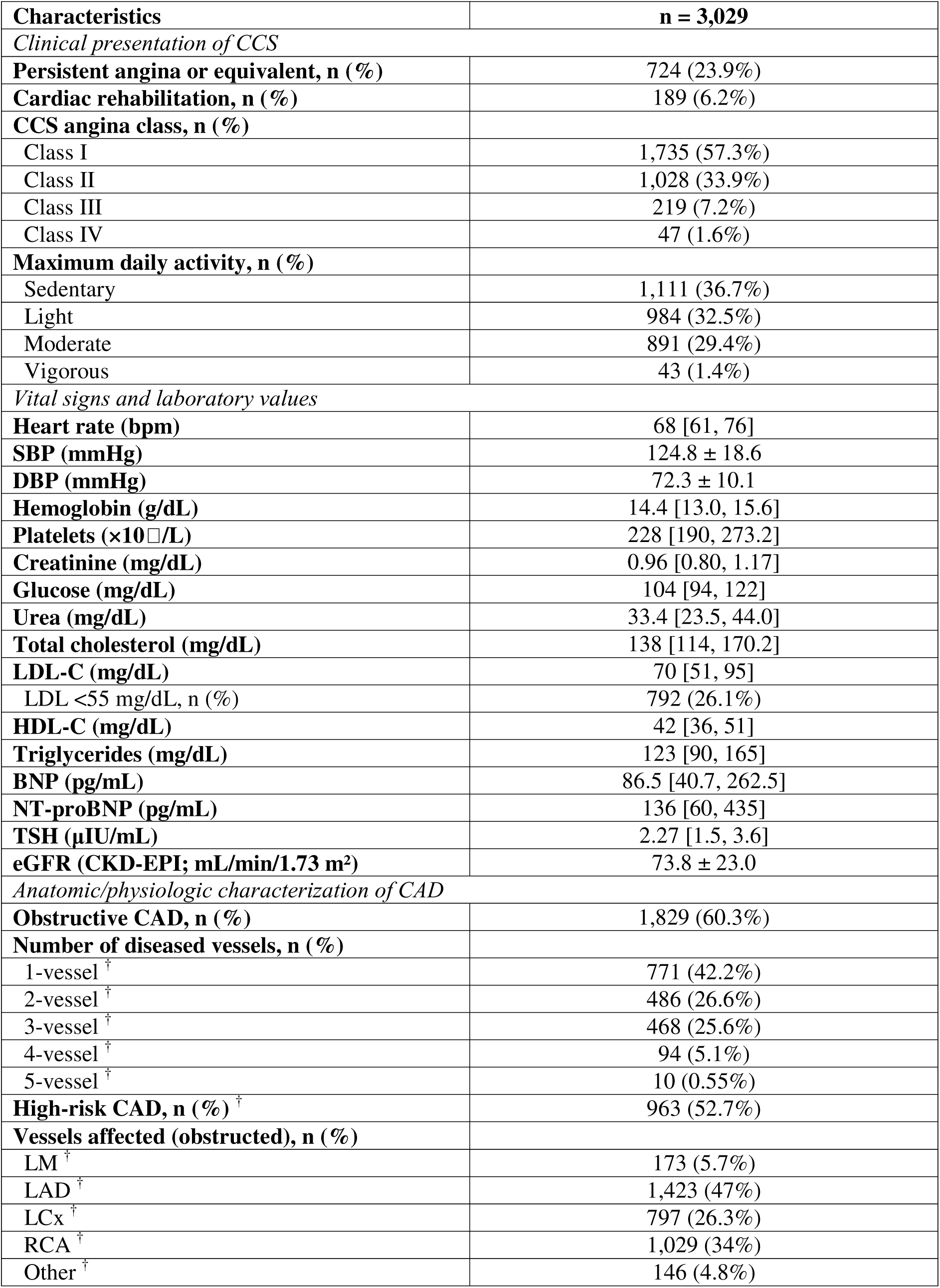
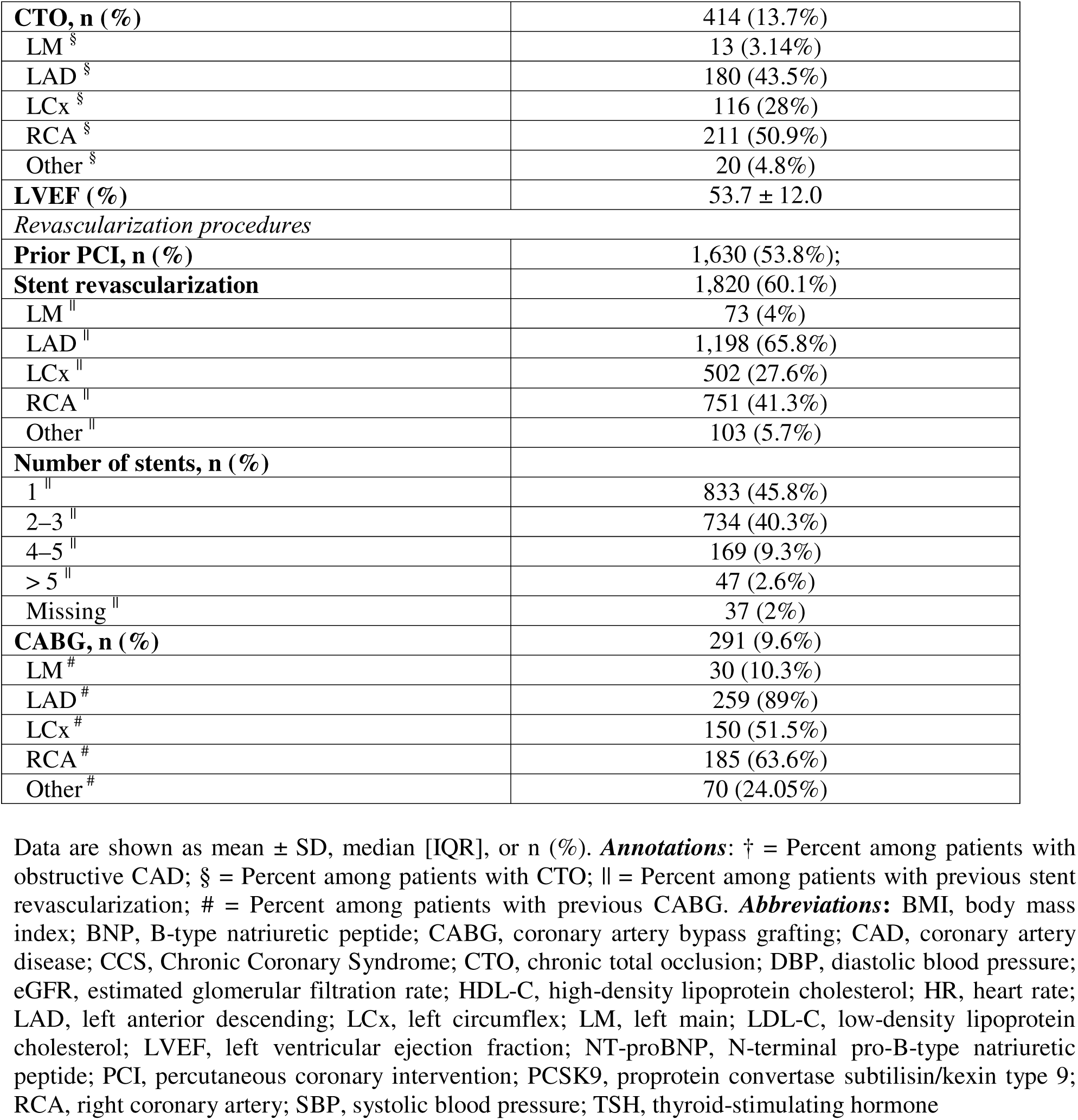
Clinical presentation, imaging findings, and procedures of the patients included in RESINCCRO.

### Anatomic characterization of CAD

Obstructive CAD was present in 60.3%, and 52.7% of those with obstruction met high-risk criteria according to the ESC definition (**Supplementary Table 4**). Among obstructive cases, disease extent was 1-vessel 42.2%, 2-vessel 26.6%, 3-vessel 25.6%, 4-vessel 5.1%, and 5-vessel 0.55% (**Figure 3A**). The most frequently involved coronary arteries were the Left Anterior Descending (LAD) (47%) and Right Coronary Artery (RCA) (34%), followed by the Left Circumflex (LCx) (26.3%) and the Left Main (LM) artery (5.7%) (**Figure 3B**). Chronic total occlusion (CTO) was identified in 13.7%, most commonly in the RCA (50.9%) and LAD (43.5%), with fewer in the LCx (28%), LM (3.14%), and other segments (4.8%) (**Figure 3C**). The mean LVEF was 53.7 (± 12.0%). Regarding revascularization history, prior PCI was performed in 53.8% and stent revascularization in 60.1%. Stents were most often placed in the LAD (65.8%) and RCA (41.3%), with additional placements in the LCx (27.6%) and LM (4%). The number of stents per patient clustered at one (45.8%) and two to three (40.3%), with fewer having four to five (9.3%) or >5 (2.6%) (**Figure 3D**). CABG had been performed in 9.6% patients, with grafts most commonly to the LAD (89%) and RCA (63.9%) (**Table 3**)

**Figure 3:**
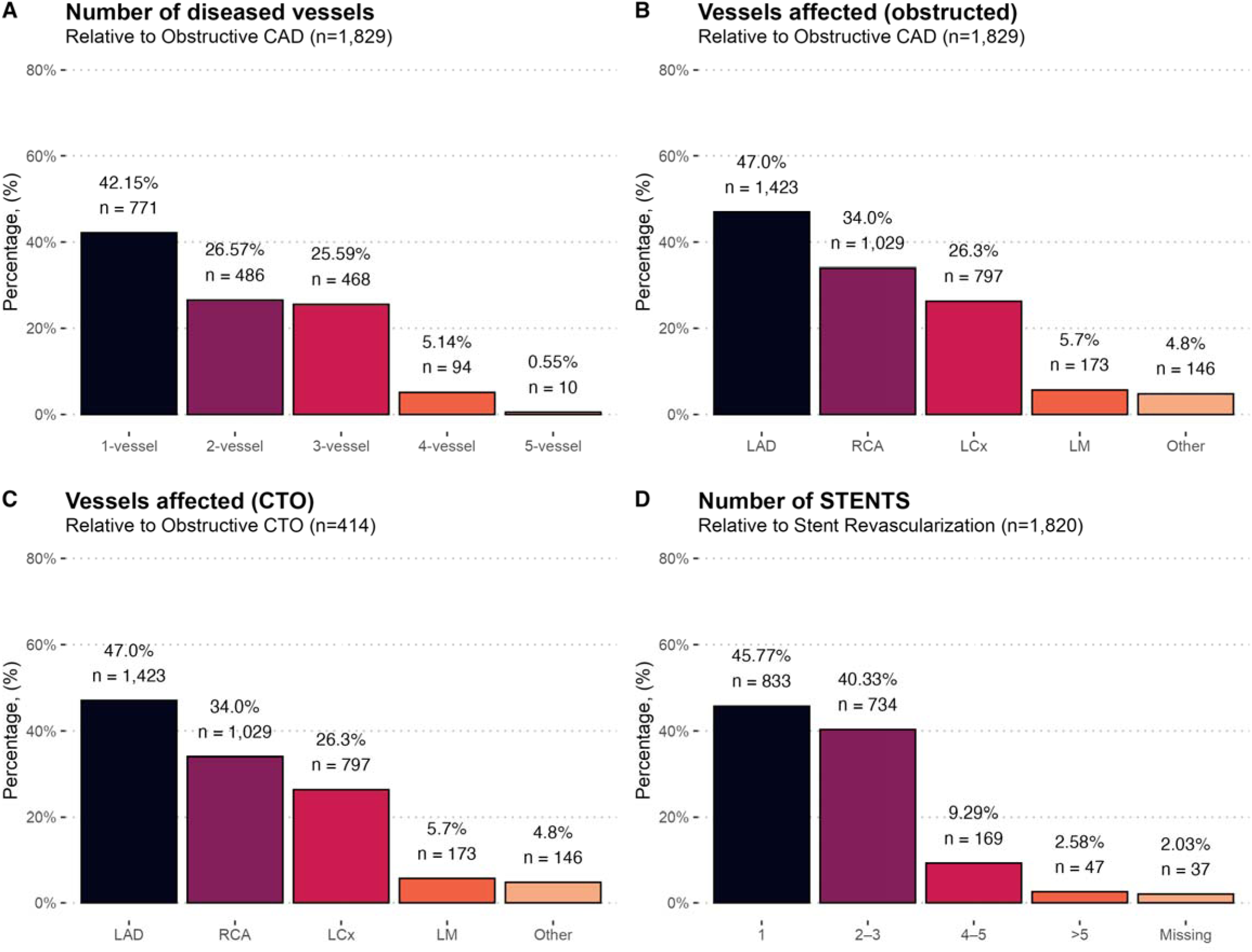
Anatomical and procedural characterization of obstructive CAD in RESINCCRO Data are shown as percentages (%) and absolute frequencies (n). *Abbreviations*: CABG, coronary artery bypass grafting; CAD, coronary artery disease; CCS, Chronic Coronary Syndrome; CTO, chronic total occlusion; LAD, left anterior descending; LCx, left circumflex; LM, left main; PCI, percutaneous coronary intervention; RCA, right coronary artery.

### Pharmacological treatments profile

Reported medications are presented in **Supplementary Table 5**. Briefly, most patients were treated with aspirin (74.6%) and clopidogrel (47.7%), whereas ticagrelor and prasugrel were rarely used. DOACs were reported in 10.6%, the majority with agents such as rivaroxaban (55.6%) and apixaban (36.9%) (**Figure 4A**). Beta-blockers were prescribed in 63.6%, primarily metoprolol (48.9%) and bisoprolol (33.4%). Among renin–angiotensin system inhibitors, ACE inhibitors were used in 19.2% of patients, most frequently enalapril (71.1%), while ARBs were more common (46.0%), being telmisartan (30.4%) and losartan (29.0%) the most used. Calcium-channel blockers were reported in one quarter of patients, with amlodipine as the predominant agent (69.7%) (**Figure 4B**). Regarding lipid-lowering therapy, statins were nearly prescribed to all patients (93.2%), most frequently atorvastatin (79.8%) and rosuvastatin (15.7%), with ezetimibe prescribed in 24.6% and PCSK9 inhibitors in 2.4% (**Figure 4C**). Antianginal/anti-ischemic medication included nitrates in 17.3% (mostly long-acting forms), trimetazidine in 10.0%, and ivabradine in 2.7% (**Figure 4D**).

**Figure 4:**
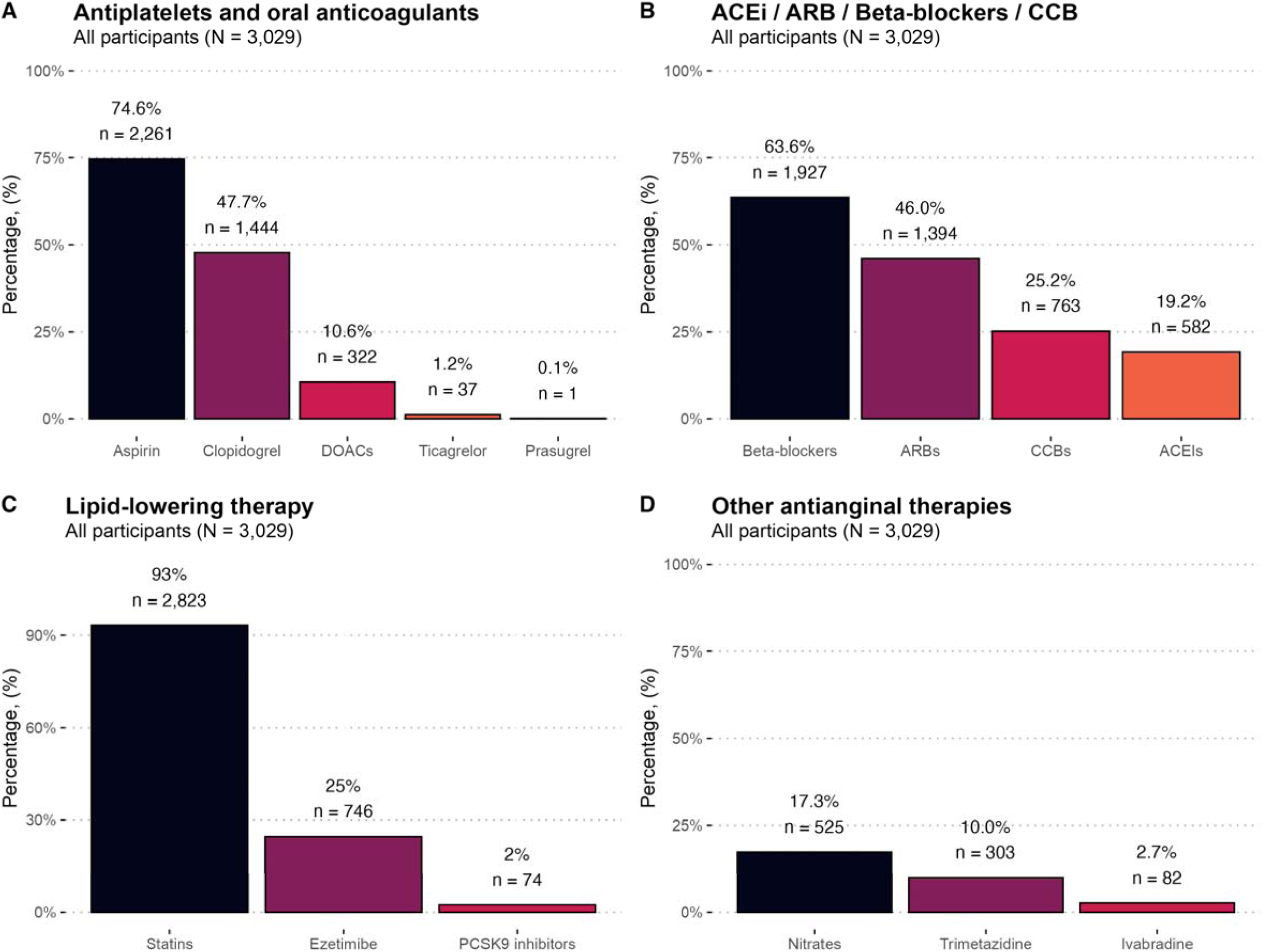
Pharmacological therapy in RESINCCRO. Data are shown as percentages (%) and absolute frequencies (n). *Abbreviations*: ACEi, angiotensin-converting enzyme inhibitors; ARB, angiotensin receptor blockers; CCB, calcium-channel blockers; DOAC, direct oral anticoagulants; PCSK9, proprotein convertase subtilisin/kexin type 9.

## DISCUSSION

In this first report of RESINCCRO, a Mexican national-level registry of adults with CCS, we aimed to describe their clinical presentation, cardiovascular risk factors, coronary anatomic characteristics, and pharmacological treatment profiles. We found a heterogeneous spectrum of coronary presentations coexisting with a high burden of cardiometabolic comorbidities. Overall, RESINCCRO highlights the complex scenario for Mexican patients living with CCS; however, our findings underscore several areas of opportunities to improve patient care in this population, such as strengthen secondary prevention, refine anti-ischemic strategies, and broaden access to cardiac rehabilitation. The real-world evidence derived from our registry may inform clinical practice, ultimately aiming to reduce the long-term burden of CCS in our country.

Our results support the view that CCS is a heterogeneous and progressive condition rather than a single, stable state. Almost one in four patients in our registry reported persistent angina or an equivalent, with symptom burden concentrated in milder classes. This pattern has also been reported in the CLARIFY registry, where 20–25% of stable CCS patients with CAD report some degree of angina (23,24). Consequently, a substantial proportion (∼75%) may have absent symptoms, which can mask the reduced coronary flow in these patients. Additionally, ∼60% of our patients had obstructive epicardial disease, implying that ∼40% had no flow-limiting stenosis. This is consistent with the Ischemia with no Obstructive Coronary Arteries (INOCA) and with Angina with no Obstructive Coronary Arteries (ANOCA) phenomenon (25). For example, in an analysis from the American College of Cardiology National Cardiovascular Data Registry made by Patel *et al.*, only 37.6% of ∼400,000 catheterized patients had obstructive CAD, implying that ∼62% lacked obstructive lesions. Moreover, in a subanalysis of the ISCHEMIA trial by Reynolds *et al.*, they demonstrated that even among CCS patients with moderate to severe ischemia, up to ∼13% had INOCA (26,27). This is relevant given that CCS patients with angina, with and without prior myocardial infarction MI, had higher rates of incident cardiovascular events and all-cause mortality, as demonstrated by Sobbert *et al.* in a subanalysis of the CLARIFY registry (28). These findings are supported by a evidence suggesting that CCS, particularly INOCA, may be driven by either vascular or microvascular dysfunction and epicardial or microvascular vasospasm which can only be identified by a acetylcholine/ergonovine test induction or a diffuse atherosclerosis detected by FFR pullback using advances intracoronary imaging techniques (29,30). These conditions are often undiagnosed, and therefore, no tailored therapy is prescribed for these patients. As consequence, these patients continue to experience recurrent angina with poor quality of life, leading to repeated hospitalizations, unnecessary repeat coronary angiography, and adverse cardiovascular outcomes (9). This evidence derived into the 2019 ESC framework, which explicitly includes vasospastic/microvascular angina and post-event states as specific subgroups of CCS which may be at higher risk of developing adverse outcomes. However, whether intensifying medical therapy, implementing antianginal/anti-ischemic medication, or implementing novel agents (e.g., zibotentan) improves outcomes remains an area for further research, aligned with ongoing RCTs such as WARRIOR and PRIZE that test mechanism-directed strategies to improve symptoms (31,32).

Our results also show that RESINCCRO patients had a high prevalence of concomitant cardiovascular risk factors (arterial hypertension, T2D, CKD, and excess adiposity) and low achievements of LDL-C goals, despite being a secondary-prevention population. Despite near-universal statin use (93.2%), the median LDL-C was 70 mg/dL, indicating that at least half of patients remain above ESC guidelines recommended for LDL-C goals (33). These findings have been reported in other international CCS cohorts. For example, in the CLARIFY registry, Steg et al. reported a high prevalence of concomitant cardiovascular risk factors among patients with stable CAD (hypertension 79%, T2D 29%) and frequent under-attainment of lipid goals (24). In the CORONOR registry, Lemesle *et al.* observed a similar high prevalence of concomitant comorbidities (T2D 31%, hypertension 60%) and an average LDL-C of 89 ± 28 mg/dL despite widespread prescription of secondary-prevention therapy (statins 92.2%, renin–angiotensin system blockers 81.9%, β-blockers 79.4%) (34). Consistent with these patterns, the EURECA registry showed that only 56% of patients fully met the 2019 ESC CCS diagnostic-pathway criteria (35). Likewise, EUROASPIRE-V reported that 70% of patients with coronary syndromes (acute and chronic) did not achieve LDL-C <70 mg/dL across 131 centers in 27 European countries (36). Finally, at a national level, our results align with RENASICA, where Martínez-Sánchez *et al.* documented high rates of hypertension, diabetes, dyslipidemia, and obesity among Mexican patients with CAD (16). Taken together, these data show that CCS patients commonly have multiple concomitant risk factors and struggle to reach lipid targets. This is often accompanied by very low use of cardiac rehabilitation (6.2%) and uneven antianginal/anti-ischemic medication optimization, which reveals implementation gaps and the need to intensify cardiometabolic control alongside mechanism-directed anti-ischemic care. Beyond chronic comorbidities, revascularization should not be interpreted as the sole strategy to relieve angina in these patients. Many patients in RESINCCRO had a long-standing history of CCS, potentially reflecting a chronic symptom burden accompanied by incomplete therapeutic optimization and delayed achievement of treatment goals. In this setting, revascularization may become one of the last resources in treatment management rather than the initial step in care. Therefore, antianginal therapy, lipid-lowering treatment, cardiometabolic risk-factor control, and cardiac rehabilitation should be intensified both before and after considering revascularization in this population (37).

Taken together, evidence from this initial report from RESINCCRO yields five clinical messages to strengthen CCS care in Mexico: (1) improve and individualize optimal medical therapy taking into account antianginal/anti-ischemic medication with timely lipid-lowering intensification beyond statins when indicated, delivered within multidisciplinary teams that address coexisting cardiometabolic conditions; (2) build and scale cardiac rehabilitation programs with explicit health-status and functional goals and routine measurement; (3) recognize that persistent angina may occur despite prior revascularization, which may yield the coexistence of multiple causes of myocardial ischemia and the importance of optimizing medical therapy even after PCI; in patients with persistent angina despite optimal medical therapy, use mechanism-directed testing (noninvasive and invasive physiology) and offer physiology-guided revascularization when ischemia is demonstrated, particularly in those with CTO; (4) there is a need to improve functional class assessment beyond traditional scales (such as the Canadian Cardiovascular Society and NYHA), for which a guided qualitative and patient-centered evaluation is needed; and (5) there is need to create registries like RESINCCRO among Latin America countries that could help define local epidemiology, identify implementation gaps, and guide resource allocation (e.g., access to non-statin lipid-lowering therapy, CR capacity, imaging infrastructure), enabling resource planning and continuous quality improvement in CCS.

### Strengths and Limitations

Our study has both strengths and limitations that should be acknowledged. Regarding strengths, RESINCCRO is the first dedicated Mexican registry focused on CCS across ∼50 centers and five regions, and the second in Latin America, to comprehensively include individual-level data on this condition (38). The inclusion of information on presentation, anatomy/physiology, revascularization history, and detailed pharmacotherapy enabled a multicenter characterization of real-world CCS care in our country. Moreover, our recruitment fulfilled an operational sample of 3,029 adults. Therefore, our estimated sample provides adequate precision to evaluate the prevalence of CCS according to our initial sample size estimation. However, several limitations should be acknowledged. First, these initial results were intended as a cross-sectional analysis based on convenience enrollment from participating sites. This may introduce selection and referral bias toward practices at tertiary centers located in northern states and Mexico City. Nonetheless, most coronary syndrome registries in Mexico, such as the RENASICA I, II and III report a similar limitation, as most tertiary cardiovascular centers are concentrated in these areas (16–18). Second, data were recorded from medical records and entered by participating clinicians, so misclassification is possible. To mitigate this, a team member reviewed a random sample of cases to verify data interpretation. Third, clinical adjudication of ischemia mechanisms and standardized functional testing across sites were not mandated, and advanced imaging or microvascular/vasospastic assessments were variably captured, which could bias classification toward clinical judgment alone. Fourth, we included prevalent CCS cases rather than incident cases, which may have excluded more severe cases from the registry and introduced survivorship bias. Finally, the absence of longitudinal follow-up limits causal inference and outcome evaluation. RESINCCRO, however, is planned to have prospective waves to evaluate the longitudinal trajectories of these patients.

## CONCLUSION

In conclusion, RESINCCRO provides a national evaluation of adults living with CCS in Mexico. Our findings reveal a highly comorbid secondary-prevention population with frequent persistent angina and limited functional capacity, suboptimal achievement of risk-factor targets despite high statin use, and very low uptake of cardiac rehabilitation. These findings underscore an urgent need for integrated, mechanism-directed care that combines optimal medical therapy, guideline-directed revascularization, and systematic rehabilitation. Overall, RESINCCRO is a valuable platform to guide personalized, evidence-based strategies, and inform potential resource allocation and policies of care aimed at improving CCS care in Mexico and the regions.

## AUTHOR CONTRIBUTIONS

Research idea and study design: BB-EA, RI-MO

Data acquisition: R-MR, LL-AA, BB-MR, GB-LG, CL-SA, RI-JM, AG-MA, NE-PL, P-JJ, RG-FJ, DM-ED, VT-EM, PS-HA, ES-MC, CA-JR, AM-LF, P-FG, RI-MO, G-H, VV-MD, N-JP, BG-MA, BR-AE, T-MQ, NG-R, AR-E, LP-JL, HH-C, RR-RH, MV-JA, BA-Y, DA-AD, GP-AI, BB-EA, R-A, RG-E, VA-JA, RC-CR, ST-LA, F-SA, LZ-JS, CR-F, AR-E, AM-A, PL-AA, BT-FM.

Data analysis/interpretation: BB-EA

Statistical analysis: BA-Y, LP-JL, MV-JA

Manuscript drafting: BB-EA, RI-MO

Supervision or mentorship: BB-EA, RI-MO

Each author contributed important intellectual content during manuscript drafting or revision and accepted accountability for the overall work by ensuring that questions pertaining to the accuracy or integrity of any portion of the work are appropriately investigated and resolved.

## Data Availability

All data produced in the present study are available upon reasonable request to the authors

## ACKNOWLEDGMENTS

We gratefully acknowledge all the investigators and healthcare personnel from all participating cardiovascular centers who contributed patients and clinical data to the RESINCCRO Mexican Registry of Chronic Coronary Syndrome.

## ABBREVIATIONS

ACS: Acute Coronary Syndromes
BMI: Body Mass Index
CAD: Coronary Artery Disease
CCS: Chronic coronary syndromes
DBP: Diastolic blood pressure
ESC: European Society of Cardiology
FFR: Fractional Flow Reserve
LAD: Left Anterior Descending Artery
LCx: Left Circumflex Artery
LM: Left Main Artery
LDL-C: Low-density lipoprotein cholesterol
RESINCCRO: REgistro de SÍNdromes Coronarios CRÓnicos (Chronic Coronary Syndromes Mexican Registry)
RCA: Right Coronary Artery
T2D: Type 2 diabetes
SBP: Systolic blood pressure

