## Supplementary Material for "Chronic Coronary Syndrome in Mexico: Design and Initial Insights from the RESINCCRO Mexican Registry"

### Supplementary Methods

*Sample size estimation*

We planned that the RESINCCRO would require a sample size to estimate sex-specific prevalence of CCS with a fixed absolute precision (half-width of the 95% CI) of **i = 0.01** (±1 percentage point). We used a single-proportion formula based on the Cochran approach (1). The precision-based sample size is:

$$N = \frac{Z^2 * P(1-P)}{i^2}$$

where $Z^2$ is the two-sided standard normal deviate (≈1.99 for 95%), $P$ is the expected prevalence, and $i$ is the tolerated half-width of the CI (precision target). We assumed a prevalence of CCS in women of 3.5% and in men of 4.3% based on the study of Bonet-Basiero et al.(2).

Thus, the sex-stratified requirement estimation could be expressed as follows:

Women:

$$N = \frac{1.99^2 * 0.035(1-0.035)}{0.01^2} = 1{,}337$$

Men:

$$N = \frac{1.99^2 * 0.043(1-0.043)}{0.01^2} = 1{,}630$$

Thus, the overall planned sample was 2,967. Our planned sample meets the target precision for both sexes and provides cushion for minor data loss and any modest center-level clustering. However, our recruitment fulfilled an operational sample of 3,029 adults. Therefore, our estimated sample provides adequate precision to evaluate the prevalence of CCS under this scenario

Supplementary Figure 1

Research and Ethics Committee Approval of the RESINCCRO (Spanish Version).

MédicaSur

Ciudad de México a 19 febrero 2025

Folio: CEI-000005
Protocolo: IC4-06795-011-MEX
Fecha de sesión: 14 febrero 2025
Sesión: Extraordinaria
Acta: CEI-233 | CI-231 | CB-227
Código Interno: 2023-EXT-793

**Enrique Alexander Berrios Barcenas**
Investigador Principal
Centro para el Desarrollo de la Medicina y de Asistencia Médica Especializada S.C
Río Choix No. 922, Col. Gral. Antonio Rosales Flores., Culiacán, Sinaloa, México., CP. C.P. 80230
P R E S E N T E

Asunto: **Re-aprobación anual**

**IC4-06795-011-MEX**
**RESINCCRO-CCSR: REgistro de SÍNdromes Coronarios CRÓnicos (Chronic Coronary Syndrome Mexican Register) 3**
**RESINCCRO**

---

Con relación al protocolo de investigación arriba mencionado, después de revisar la documentación presentada, le informo que el Comité de Ética en Investigación de Médica Sur, S.A.B. de C.V., ha decidido otorgar la re-aprobación anual del protocolo.

Le informo que de acuerdo al riesgo que representa el presente proyecto, debe enviar a este Comité un informe de avances de manera anual, y deberá solicitar la renovación anual del presente dictamen, **un mes antes** de la fecha de expiración del mismo, el cual tiene una vigencia a **Febrero 2026**.

Siendo el quórum requerido para la validez de este Dictamen, cabe mencionar que los miembros que participaron en la revisión fueron:

| Nombre | Profesión \| Disciplina | Puesto |
|---|---|---|
| Norberto Carlos Chávez Tapia | Médico Especialista | Presidente |
| Karen Dennis Garcia López | Abogada | Vocal Secretario |
| Alba Cicero Casarrubias | Médico Especialista | Vocal |
| Álvaro Lomelí Rivas | Médico Especialista | Vocal |
| David Francisco Cantú De León | Médico Especialista | Vocal |
| Noemí Santos Caballero | Médico Especialista | Vocal |
| Oscar Daniel Mendoza Saavedra | QFB | Vocal |
| Jorge Guillermo Chávez Abraján | Abogado | Representante del Núcleo Afectado |

---

Supplementary Table 1

Geographical distribution of patients by region and state included in the RESINCCRO registry.

| **Region †<br>Frequency (%)** | **State** | **Included patients<br>N = 3,029** |
|---|---|---|
| Central<br>n = 109 (3.6%) | State of Mexico | 10 (0.3%) |
| | Hidalgo | 43 (1.4%) |
| | Morelos | 51 (1.7%) |
| | Puebla | 5 (0.2%) |
| Central west<br>n = 570 (18.8%) | Aguascalientes | 28 (0.9%) |
| | Guanajuato | 68 (2.2%) |
| | Jalisco | 65 (2.1%) |
| | Michoacán | 1 (0.0%) |
| | Querétaro | 304 (10.0%) |
| | San Luis Potosí | 80 (2.6%) |
| | Zacatecas | 24 (0.8%) |
| Mexico City<br>n = 732 (24.2%) | Mexico City | 732 (24.2%) |
| North<br>n = 985 (32.5%) | Baja California | 39 (1.3%) |
| | Chihuahua | 23 (0.8%) |
| | Coahuila | 322 (10.6%) |
| | Durango | 5 (0.2%) |
| | Nuevo León | 31 (1.0%) |
| | Sinaloa | 335 (11.1%) |
| | Sonora | 224 (7.4%) |
| | Tamaulipas | 6 (0.2%) |
| South<br>n = 633 (20.9%) | Campeche | 3 (0.1%) |
| | Chiapas | 5 (0.2%) |
| | Guerrero | 22 (0.7%) |
| | Oaxaca | 10 (0.3%) |
| | Quintana Roo | 2 (0.1%) |
| | Tabasco | 112 (3.7%) |
| | Veracruz | 4 (0.1%) |
| | Yucatán | 475 (15.7%) |

Data are shown as n (%).
*Footnotes*: † = Region rows display the subtotal for each region; state rows present the breakdown within that region.

Supplementary Table 2

Description and definitions of the eligibility criteria to be included in the RESINCCRO

| |
|---|
| **Inclusion criteria** |
| Adults ≥18 years of any sex, able and willing to provide informed consent and with at least one of the following criteria: |
| 1) Stable angina meeting ≥1 typical feature and lasting ≤10 minutes: a. (i) oppressive discomfort in the anterior chest, neck, jaw, shoulder, or arm b. (ii) precipitated by physical exertion c. (iii) relieved by rest or sublingual nitrates within <5 minutes. [a] |
| 2) Angina equivalents with documented ischemia on non-invasive stress testing. [b] |
| 3) Prior CCS diagnosis based on clinical evaluation and/or non-invasive or invasive imaging consistent with ESC criteria. |
| 4) Previously detected anatomically obstructive CAD on imaging (stenosis >50% or FFR ≤0.80). |
| 5) Local expert diagnosis of ischemic heart disease / coronary artery disease. |
| 6) Signs or symptoms of heart failure plus ≥1 criterion above. [d] |
| **Exclusion Criteria** |
| Acute coronary syndrome within the previous 2 months |
| Unstable angina within the previous 3 months |
| Severe valvular heart disease |
| Storage or infiltrative cardiomyopathy |
| Hypertrophic, dilated, or hereditary cardiomyopathy |
| Aortic disease |
| Pericardial disease or myocarditis |
| Stress-induced (Takotsubo) cardiomyopathy |
| Pulmonary hypertension |
| Cardiac tumors |
| Congenital heart disease |
| Active or terminal cancer |
| Positive pregnancy test |
| **Elimination criteria** |
| Inability to comply with the protocol or refusal to participate (withdrawal of consent) |
| Incomplete medical records |

**Abbreviations:** CCS, chronic coronary syndrome; CAD, coronary artery disease; ESC, European Society of Cardiology; FFR, fractional flow reserve; ECG, electrocardiogram; CMR, cardiovascular magnetic resonance; SPECT, single-photon emission computed tomography; PET, positron emission tomography; NYHA, New York Heart Association; LVEF, left ventricular ejection fraction.

**Footnotes:**

a) Stable angina definition mirrors classical features; capture Canadian Cardiovascular Society class at enrollment.
b) Stress modalities may include exercise ECG, stress echocardiography, SPECT/PET perfusion imaging, or stress CMR; record modality and result (ischemia present/absent; extent if available).
c) Align with 2019 ESC guidance for CCS (3).
d) Symptoms/signs per NYHA and clinical exam; document natriuretic peptides and LVEF when available.

Supplementary Table 3

STROBE cross-sectional guidelines report for study. This checklist was completed on February 18th, 2026 using https://www.goodreports.org/ a tool made by the EQUATOR Network in collaboration with Penelope.ai (von Elm E, Altman DG, Egger M, Pocock SJ, Gotzsche PC, Vandenbroucke JP. The Strengthening the Reporting of Observational Studies in Epidemiology (STROBE) Statement: guidelines for reporting observational studies)

| | | Reporting Item | Page Number |
|---|---|---|---|
| **Title and abstract** | | | |
| Title | #1a | Indicate the study's design with a commonly used term in the title or the abstract | 1 |
| Abstract | #1b | Provide in the abstract an informative and balanced summary of what was done and what was found | 5 |
| **Introduction** | | | |
| Background / rationale | #2 | Explain the scientific background and rationale for the investigation being reported | 7-8 |
| Objectives | #3 | State specific objectives, including any prespecified hypotheses | 8 |
| **Methods** | | | |
| Study design | #4 | Present key elements of study design early in the paper | 8 |
| Setting | #5 | Describe the setting, locations, and relevant dates, including periods of recruitment, exposure, follow-up, and data collection | 8-9 |
| Eligibility criteria | #6a | Give the eligibility criteria, and the sources and methods of selection of participants. | 9 |
| | #7 | Clearly define all outcomes, exposures, predictors, potential confounders, and effect modifiers. Give diagnostic criteria, if applicable | 9 |
| Data sources / measurement | #8 | For each variable of interest give sources of data and details of methods of assessment (measurement). Describe comparability of assessment methods if there is more than one group. Give information separately for for exposed and unexposed groups if applicable. | 9-10 |
| Bias | #9 | Describe any efforts to address potential sources of bias | N/A |
| Study size | #10 | Explain how the study size was arrived at | 10 |
| Quantitative variables | #11 | Explain how quantitative variables were handled in the analyses. If applicable, describe which groupings were chosen, and why | N/A |
| Statistical methods | #12a | Describe all statistical methods, including those used to control for confounding | 10 |
| Statistical methods | #12b | Describe any methods used to examine subgroups and interactions | N/A |
| Statistical methods | #12c | Explain how missing data were addressed | 10 |
| Statistical methods | #12d | If applicable, describe analytical methods taking account of sampling strategy | 10 |
| Statistical methods | #12e | Describe any sensitivity analyses | N/A |
| **Results** | | | |

| | | | |
|---|---|---|---|
| Participants | #13a | Report numbers of individuals at each stage of study—eg numbers potentially eligible, examined for eligibility, confirmed eligible, included in the study, completing follow-up, and analysed. Give information separately for for exposed and unexposed groups if applicable. | 10,11,12 |
| Participants | #13b | Give reasons for non-participation at each stage | N/A |
| Participants | #13c | Consider use of a flow diagram | N/A |
| Descriptive data | #14a | Give characteristics of study participants (eg demographic, clinical, social) and information on exposures and potential confounders. Give information separately for exposed and unexposed groups if applicable. | 10 |
| Descriptive data | #14b | Indicate number of participants with missing data for each variable of interest | ST4 |
| Outcome data | #15 | Report numbers of outcome events or summary measures. Give information separately for exposed and unexposed groups if applicable. | 10,11 |
| Main results | #16a | Give unadjusted estimates and, if applicable, confounder-adjusted estimates and their precision (eg, 95% confidence interval). Make clear which confounders were adjusted for and why they were included | N/A |
| Main results | #16b | Report category boundaries when continuous variables were categorized | 10,11,12 |
| Main results | #16c | If relevant, consider translating estimates of relative risk into absolute risk for a meaningful time period | N/A |
| Other analyses | #17 | Report other analyses done—e.g., analyses of subgroups and interactions, and sensitivity analyses | N/A |
| **Discussion** | | | |
| Key results | #18 | Summarise key results with reference to study objectives | 12,13 |
| Limitations | #19 | Discuss limitations of the study, taking into account sources of potential bias or imprecision. Discuss both direction and magnitude of any potential bias. | 19,20 |
| Interpretation | #20 | Give a cautious overall interpretation considering objectives, limitations, multiplicity of analyses, results from similar studies, and other relevant evidence. | 13,14,15 |
| Generalisability | #21 | Discuss the generalisability (external validity) of the study results | 14,15 |
| **Other Information** | | | |
| Funding | #22 | Give the source of funding and the role of the funders for the present study and, if applicable, for the original study on which the present article is based | 17 |

Supplementary Table 4

Conceptual, operational definitions, units, and missing information of the assessed variables from the RESINCCRO.

| Variable | Brief definition | Unit of measurement | Missing Values |
|---|---|---|---|
| **Demographic and Socioeconomic variables** | | | |
| Center code | Pre-assigned code for recruiting center | Text | 0% |
| Patient code | Center code + pre-assigned patient code | Text | 0% |
| Recruitment date | Day/Month/Year recorded in medical record | Date (YYYY-MM-DD) | 0% |
| Age | Age in years according to medical record | Years | 0% |
| Sex | Sex registered in medical record | Male / Female | 0% |
| Institution | Facility name recorded in medical record | Public / Social security / Private | 0% |
| Insurance | Governmental or insurance plan recorded in file | Yes/No | 0% |
| Place of birth | State or country from medical record | Text | 0% |
| Place of residence | Urban/rural and municipality from medical record | Text + Urban/Rural | 0% |
| Attending specialty | Specialty of attending physician | Cardiology / Internal medicine / Other | 0% |
| **Cardiovascular risk factors and self-reported concomitant comorbidities** | | | |
| BMI | Weight (kg) / height$^2$ (mt$^2$) | kg/m$^2$ | 0% |
| BMI categories | Derived from BMI | Underweight <18.5; Normal 18.5–24.9; Overweight 25.0–29.9; Obesity I 30.0–34.9; Obesity II 35.0–39.9; Obesity III ≥40 | 0% |
| Arterial hypertension | Clinical record | Yes/No | <1% |
| Type 2 diabetes | Clinical record | Yes/No; specify complications (nephropathy/retinopathy/neuropathy) | <1% |
| Thyroid dysfunction | Hyperthyroidism: TSH <0.1 µIU/mL with high T4; Hypothyroidism: TSH ≥20 µIU/mL with low T4, per lab reference | Hyper / Hypo / None | 0% |
| Chronic kidney disease | eGFR <60 mL/min/1.73m² for ≥3 months (CKD-EPI 2021); record G1–G5 stage | Yes/No; Stage G1–G5 | 3.9% |
| COPD | Clinical record | Yes/No | <1% |
| Anemia | Hb <12 g/dL (women) or <14 g/dL (men), or Hct <36% (women) / <41% (men) | Yes/No | 7.1% |

| | | | |
|---|---|---|---|
| Chronic liver disease | Clinical history | Yes/No | <1% |
| GERD | Clinical and Endoscopic diagnosis | Yes/No | 1.1% |
| Osteoporosis | DXA or clinical diagnosis | Yes/No | 14.4% |
| Smoking | Current daily or occasional | Yes/No | 0% |
| Alcohol use disorder | Clinical history | Yes/No | 0% |
| Coronary artery disease | Prior angiographically confirmed CAD | Yes/No | 0% |
| Peripheral arterial disease | Clinical record | Yes/No | <1% |
| Carotid artery disease | Ultrasound/CT/MRI documented carotid atherosclerosis | Yes/No | 7.7% |
| Cerebrovascular events | Clinical record | Yes/No; Ischemic/Hemorrhagic | <1% |
| Transient ischemic attack | Clinical record | Yes/No | <1% |
| Atrial fibrillation | Clinical record | Yes/No | <1% |
| Sinus node dysfunction | Clinical diagnosis or EP study | Yes/No | 0% |
| Venous thromboembolism | Ultrasound/CT evidence in record | Yes/No | <1% |
| **Clinical presentation of CCS** | | | |
| Persistent angina / equivalent | Investigator-reported presence of persistent angina or equivalents | Yes/No | <1% |
| CCS angina class | Investigator-graded CCS class at enrollment | Class I/II/III/IV | 0% |
| Maximum daily activity | Highest usual activity level the patient can perform without chest pain or significant symptoms, per standardized categories | I: Sedentary (sitting/lying/TV); II: Light (slow walking, light chores); III: Moderate (brisk walking, household tasks, cycling); IV: Vigorous (jog/run, tennis, swimming) | 0% |
| **Imaging studies and anatomic/physiologic characterization of CAD** | | | |
| Obstructive CAD | ≥70% in any epicardial vessel or ≥50% in left main on ICA/CCTA | Yes/No | 4.5% |
| Vessels affected (names) | LM, LAD, LCx, RCA, other (specify) | Categorical (multi-select) | 0% |
| Number of vessels affected | Count of vessels with ≥70% stenosis (or ≥50% LM) | 0/1/2/3 (±LM) | 0% |
| High-risk CAD | Based on the 2019-ESC guidelines as: Left main ≥50% OR ≥70% in 3 vessels OR ≥70% in 2 vessels including proximal LAD | Yes/No | 3.3% |

| | | | |
|---|---|---|---|
| Chronic total occlusion (CTO) | Diagnosis of CTO | Yes/No | 5.3% |
| CTO vessels affected | LM, LAD, LCx, RCA, other (specify) | Categorical (multi-select) | 0% |
| Left ventricular ejection fraction (LVEF) | Most recent LVEF by echo/CMR/nuclear | Percent (%) | 0% |
| **Procedures** | | | |
| Prior PCI | Any prior PCI recorded in medical file | Yes/No | <1% |
| Prior CABG | CABG for obstructive CAD | Yes/No | <1% |
| Stents placed (number) | Number recorded in file | Count | <1% |
| **Medications (current/previous)** | | | |
| Aspirin | Use recorded | Yes/No | 0% |
| Clopidogrel | Use recorded | Yes/No | 0% |
| Prasugrel | Use recorded | Yes/No | 0% |
| Ticagrelor | Use recorded | Yes/No | 0% |
| Direct oral anticoagulant (DOAC) | Dabigatran/rivaroxaban/apixaban/edoxaban/betrixaban use | Yes/No | 0% |
| Beta-blocker | Atenolol, bisoprolol, metoprolol, propranolol, etc. | Yes/No | 0% |
| ACE inhibitor | Captopril, enalapril, lisinopril, etc. | Yes/No | 0% |
| ARB | Losartan, valsartan, candesartan, etc. | Yes/No | 0% |
| Statin | Atorvastatin/rosuvastatin/etc. | Yes/No | 0% |
| Ezetimibe | Use recorded | Yes/No | 0% |
| PCSK9 inhibitor | Alirocumab / Evolocumab | Yes/No | 0% |
| Nitrates | GTN/dinitrate/mononitrate of isosorbide | Yes/No | 0% |
| Trimetazidine | Use recorded | Yes/No | 0% |
| Ivabradine | Use recorded | Yes/No | 0% |

***Abbreviations**:* ACE, angiotensin-converting enzyme; ARB, angiotensin II receptor blocker; BMI, body mass index; CABG, coronary artery bypass grafting; CAD, coronary artery disease; CCS, Chronic Coronary Syndrome; CCS class, Canadian Cardiovascular Society angina class; CCTA, coronary computed tomographic angiography; CKD-EPI, Chronic Kidney Disease Epidemiology Collaboration; CMR, cardiovascular magnetic resonance; COPD, chronic obstructive pulmonary disease; CT, computed tomography; CTO, chronic total occlusion; DOAC, direct oral anticoagulant; DXA, dual-energy X-ray absorptiometry; eGFR, estimated glomerular filtration rate; ECG, electrocardiogram; EP, electrophysiology; GERD, gastroesophageal reflux disease; GTN, glyceryl trinitrate; Hb, hemoglobin; Hct, hematocrit; ICA, invasive coronary angiography; LAD, left anterior descending; LCx, left circumflex; LM, left main; LVEF, left ventricular ejection fraction; MRI, magnetic resonance imaging; PCI, percutaneous coronary intervention; PCSK9, proprotein convertase subtilisin/kexin type 9; RCA, right coronary artery

Supplementary Table 5

Medication use of the patients included the RESINCCRO registry.

| Characteristics | N = 3,029 |
|---|---|
| **Aspirin, n (%)** | 2,261 (74.6%) |
| **Clopidogrel, n (%)** | 1,444 (47.7%) |
| **Ticagrelor, n (%)** | 37 (1.2%) |
| **Prasugrel, n (%)** | 1 (0.1%) |
| **Oral anticoagulants, n (%)** | 322 (10.6%) |
| Apixaban † | 119 (36.9%) |
| Dabigatran † | 8 (2.5%) |
| Rivaroxaban † | 179 (55.6%) |
| Acenocoumarol † | 15 (4.7%) |
| Warfarin † | 1 (0.1%) |
| **ACE inhibitors, n (%)** | 582 (19.2%) |
| Captopril § | 14 (2.4%) |
| Enalapril § | 414 (71.1%) |
| Lisinopril § | 28 (4.8%) |
| Perindopril § | 43 (7.4%) |
| Quinapril § | 1 (0.2%) |
| Ramipril § | 58 (10%) |
| Trandolapril § | 5 (0.9%) |
| Other § | 19 (3.3%) |
| **ARBs, n (%)** | 1,394 (46.0%) |
| Azilsartan ǁ | 20 (1.4%) |
| Candesartan ǁ | 118 (8.5%) |
| Eprosartan ǁ | 1 (0.1%) |
| Fimasartan ǁ | 13 (0.9%) |
| Irbesartan ǁ | 73 (5.2%) |
| Losartan ǁ | 404 (29%) |
| Olmesartan ǁ | 76 (5.5%) |
| Telmisartan ǁ | 424 (30.4%) |
| Valsartan ǁ | 231 (16.6%) |
| Other ǁ | 34 (2.4%) |
| **Beta-blockers, n (%)** | 1,927 (63.6%) |
| Atenolol # | 29 (1.5%) |
| Bisoprolol # | 644 (33.4%) |
| Carvedilol # | 156 (8.1%) |
| Metoprolol # | 942 (48.9%) |
| Nebivolol # | 138 (7.2%) |
| Propranolol # | 14 (0.7%) |
| Other # | 4 (0.2%) |
| **Calcium-channel blockers, n (%)** | 763 (25.2%) |
| Amlodipine †† | 532 (69.7%) |
| Diltiazem †† | 83 (10.9%) |
| Felodipine †† | 10 (1.3%) |
| Lercanidipine †† | 40 (5.2%) |
| Nifedipine †† | 87 (11.4%) |
| Verapamil †† | 11 (1.4%) |
| **Statins, n (%)** | 2,823 (93.2%) |

| | |
|---|---|
| Simvastatin §§ | 103 (3.6%) |
| Atorvastatin §§ | 2,254 (79.8%) |
| Fluvastatin §§ | 2 (0.1%) |
| Pitavastatin §§ | 2 (0.1%) |
| Pravastatin §§ | 18 (0.6%) |
| Rosuvastatin §§ | 444 (15.7%) |
| **Ezetimibe, n (%)** | 746 (24.6%) |
| **PCSK9 inhibitors, n (%)** | 74 (2.4%) |
| **Nitrates, n (%)** | 525 (17.3%) |
| Short-acting ‡ | 200 (38.1%) |
| Long-acting ‡ | 325 (61.9%) |
| **Trimetazidine, n (%)** | 303 (10.0%) |
| **Ivabradine, n (%)** | 82 (2.7%) |

Data are shown as n (%). ***Annotations***: † = Percent among patients receiving oral anticoagulants; § = Percent among patients receiving ACE inhibitors; ‖ = Percent among patients receiving ARBs; # = Percent among patients receiving beta blockers; †† = Percent among patients receiving calcium-channel blockers; §§ = Percent among patients receiving Statins; ‡ = Percent among patients receiving nitrates. ***Abbreviations*****:** ACE, angiotensin-converting enzyme; ARB, angiotensin II receptor blocker; DOAC, direct oral anticoagulant; PCSK9, proprotein convertase subtilisin/kexin type 9.
